# Automated Insomnia Phenotyping from Electronic Health Records: Leveraging Large Language Models to Decode Clinical Narratives

**DOI:** 10.1101/2025.06.02.25328701

**Authors:** Guillermo Lopez-Garcia, Davy Weissenbacher, Matthew Stadler, Karen O’Connor, Dongfang Xu, Lauren Gryboski, Jared Heavens, Noor Abu-el-Rub, Diego R. Mazzotti, Subhajit Chakravorty, Graciela Gonzalez-Hernandez

**Affiliations:** Department of Computational Biomedicine, Cedars-Sinai Medical Center, West Hollywood, CA; Perelman School of Medicine, University of Pennsylvania, Philadelphia, PA; Division of Pulmonary, Critical Care and Sleep Medicine, Department of Internal Medicine, University of Kansas Medical Center, Kansas City, KS; Division of Medical Informatics, Department of Internal Medicine, University of Kansas Medical Center, Kansas City, KS

## Abstract

Insomnia is a highly prevalent but often underdiagnosed condition in clinical practice. Its inconsistent documentation in electronic health records (EHRs) limits population-level analyses and obstructs efforts to evaluate treatment patterns or outcomes. We present a novel, fully automated approach for phenotyping insomnia directly from unstructured clinical notes using generative large language models (LLMs). Leveraging prompt engineering with few-shot learning and chain-of-thought reasoning, we evaluated our system on two distinct corpora: inpatient clinical notes from MIMIC-III and outpatient primary care notes from the University of Kansas Health System (KUMC). Our models—Llama 70B and Llama 405B—achieved F1 scores of 93.0 on the MIMIC corpus and 85.7 on the KUMC corpus, substantially outperforming domain-adapted BERT-based classifiers. Ultimately, our framework offers a scalable and interpretable solution for clinical phenotyping of insomnia and can serve as a blueprint for similar efforts targeting other underdiagnosed or under-documented conditions in the EHR.

## 1 Introduction

Insomnia is a disorder affecting about 10% of the population, causing difficulty in initiating or maintaining sleep. If left untreated, insomnia can lead to psychiatric and substance abuse disorders, psychological problems, reduced quality of life, increased absenteeism from work, and an increased risk of motor vehicle accidents, falls, and suicidal behavior. Due to its frequent co-occurrence with these conditions, primary care and mental health providers are often the initial professionals to evaluate and treat insomnia.

A notable challenge observed in clinical settings and verified in research studies is a disconnect between diagnosis and treatment of insomnia, whereby individuals are not diagnosed with insomnia in a clinical encounter, yet they are counseled or prescribed hypnotic medications commonly used to treat the disorder. Albrecht and colleagues analyzed Medicare claims data from 2006 to 2013 and found that while the diagnosis of insomnia rose from 3.9% in 2006 to 6.2% in 2013, the use of hypnotic medications increased from 21.0% to 29.6% during the same period [1]. In a retrospective analysis of Department of Veterans Affairs (VA) electronic health records (EHR) data from 2011 to 2019, Bramoweth and colleagues described that out of 439,887 Veterans in the dataset, only 6% had a diagnosis of insomnia, while 15% were prescribed hypnotic medications. They concluded that prescribing sleep medications remained the primary method of identifying Veterans with insomnia within the VA EHR [3]. Additionally, Ulmer and colleagues surveyed VA healthcare providers and found that only 39.2% of providers reported documenting insomnia diagnosis in their problem list. Reasons for this behavior included “competing diagnoses”, “lack of time”, “insomnia seeming more like a symptom than a diagnosis”, and the presence of other mental health diagnoses [22].

The lack of consistent representation of insomnia diagnosis in the EHR hinders our ability to study the characteristics of insomnia within a healthcare system, such as its clinical prevalence, risk factors, protective factors, trajectory over time, and the effectiveness of commonly prescribed medications for its treatment. As noted, the prescription of hypnotic medications has been used as a proxy for insomnia diagnosis [3]. However, this approach may underestimate the number of patients who reported symptoms that met criteria for insomnia during a clinical visit but were not offered or refused medication.

In this study, we propose leveraging Natural Language Processing (NLP) to streamline identification of insomnia from EHR clinical notes. Modern NLP systems primarily rely on artificial neural networks, with the transformer architecture being the most prevalent. Such systems split text into sequences of tokens. Tokens are then converted into vectors using a reference table. These tokens are adjusted based on their importance. Large Language Models (LLMs) based on decoder transformers deliver state-of-the-art performance, achieving remarkable results across a wide range of tasks. LLM can learn statistical relationships from vast amounts of text in a self-supervised or semi-supervised fashion.

Automatic identification of insomnia using structured and unstructured variables in the EHR was attempted by one prior study [13]. Structured variables included diagnostic codes for International Classification of Diseases 9th Revision (ICD-9) and prescriptions of hypnotic drugs. The unstructured data included keywords related to sleep and psychiatric disorders. The study showed that a combination of structured and unstructured data was optimal and that about 90% of the individuals identified with their algorithm were not diagnosed with insomnia. Though promising, the study included only ten keywords, severly limiting its contextual scope. Our proposed LLM method represents a paradigm shift in scalability, accuracy, and clinical utility, leveraging modern NLP to decode unstructured narratives at unprecedented depth for optimal identification of patients with insomnia.

Building upon prior efforts that combined structured and limited unstructured EHR data for insomnia phenotyping, our study advances the field by harnessing the power of modern large language models (LLMs) to analyze the full richness of clinical narratives. Unlike earlier approaches that relied on predefined keyword lists or manual feature engineering, our method leverages transformer-based NLP to automatically extract nuanced insomnia-related information from diverse clinical notes. Specifically, our contributions are as follows: (1) we develop and validate an LLM-based pipeline capable of identifying insomnia phenotypes from unstructured EHR data with greater contextual understanding and accuracy; (2) we demonstrate the generalizability of our approach across both the MIMIC-III and large tertiary healthcare system datasets, showing robust performance in varied clinical environments; (3) we provide a comprehensive analysis of the linguistic patterns and clinical contexts in which insomnia is documented, offering new insights into underdiagnosis and treatment trends; and (4) we make our annotated dataset and codebase publicly available to facilitate further research in computational phenotyping. Collectively, our work establishes a scalable, transferable framework for high-fidelity insomnia detection in EHRs, paving the way for improved epidemiological studies and clinical decision support.

## 2 Methods

### 2.1 Data Collection and Preparation

To develop and validate our EHR-based insomnia phenotype detection system, we curated two distinct corpora: one from the publicly available MIMIC-III dataset [11] that can be distributed to reproduce our results, and another from outpatient encounters at the University of Kansas Health System. This dual-corpus strategy enabled us to assess both the reproducibility and generalizability of our approach across diverse clinical settings.

#### MIMIC corpus

MIMIC-III is a comprehensive, de-identified database containing over 2 million clinical notes from more than 40,000 patients admitted to the intensive care units at Beth Israel Deaconess Medical Center between 2001 and 2012 [11]. For this study, we focused on the “Discharge summaries” and “Nursing/other” notes, *i*.*e*. 882,149 notes, as shown in Figure 1, as they were the most likely to mention signs and symptoms of insomnia.

**Figure 1:**
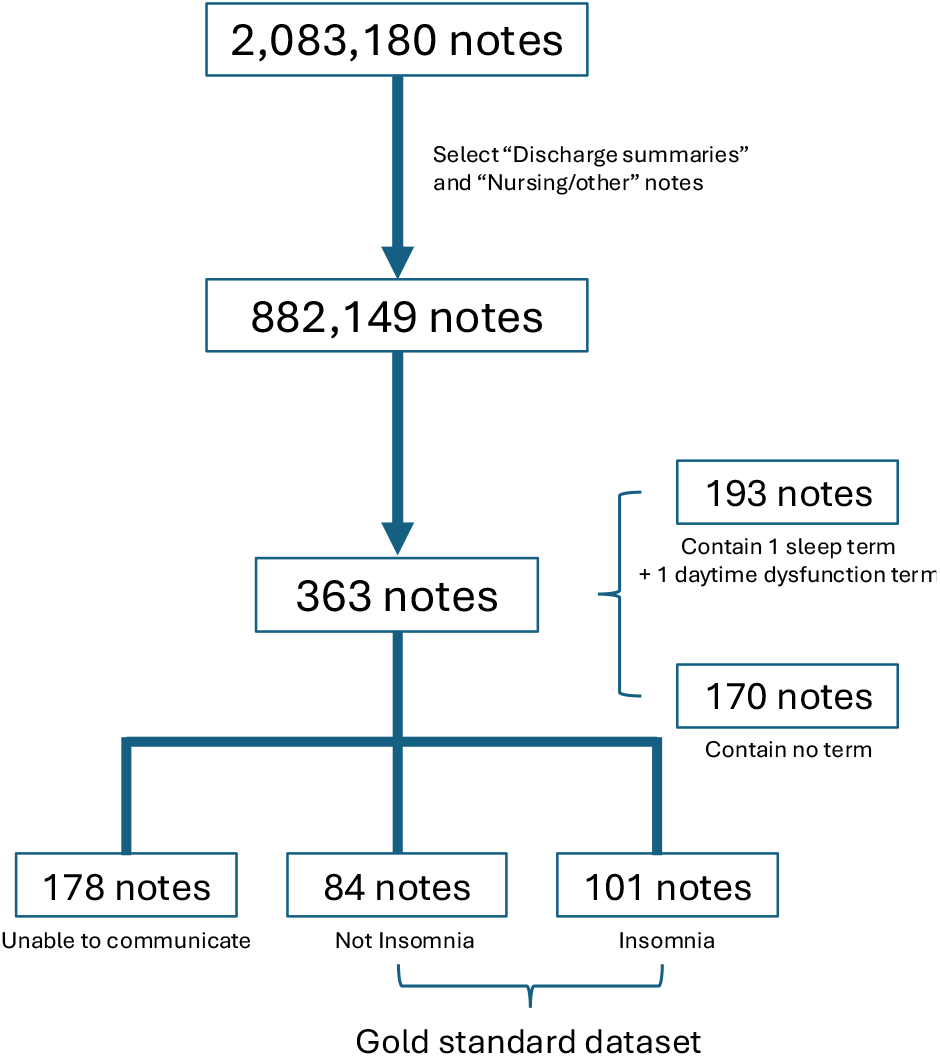
Overview of the clinical notes collection utilized for the development of our EHR-based insomnia phenotype.

We identified a subset of notes using a list of sleep-related terms manually curated by a physician (SC). The list included 118 sleep disturbance terms, such as “impaired sleep” and “frequent night waking”, as well as 337 daytime dysfunction terms, such as “drowsiness” and “fatigued” (see Supplementary Material for the full lists of terms). We randomly selected 193 notes from the “Discharge summaries” and “Nursing/other” notes that contained both sleep and daytime dysfunction terms, indicative of patients likely suffering from insomnia, and supplemented it with a subset of 170 notes that did not contain either sleep or daytime dysfunction terms, resulting in a dataset of 363 notes that we manually annotated. To ensure accurate phenotyping, from these 363 notes, we manually identified and excluded 178 notes from patients who were unable to communicate due to heavy sedation, confusion, agitation, coma, or mechanical ventilation, as their insomnia status could not be determined, leaving us with a final dataset of 185 clinical notes.

For each note, we appended available structured patient data—including age, sex, and all medications prescribed during the corresponding hospital stay— by linking notes to their corresponding hospital admissions, as illustrated in Figure 2.

**Figure 2:**
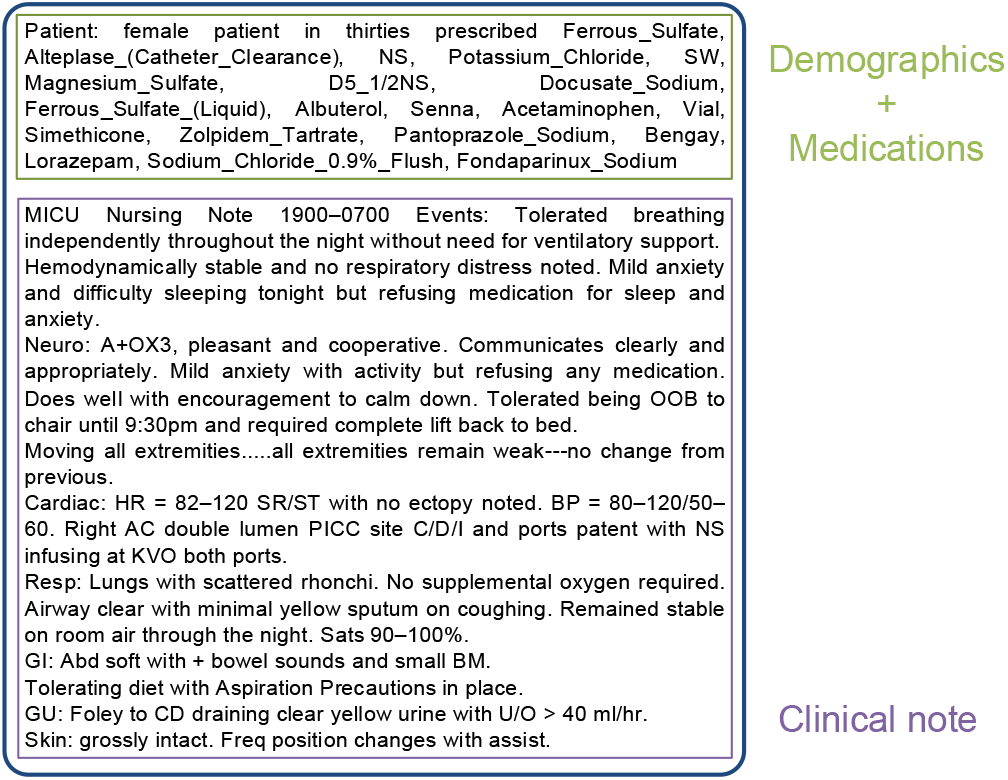
Each clinical note from MIMIC was enriched with both demographic and medication information.

#### KUMC corpus

To further evaluate model generalizability, we evaluated our approach on a second corpus from 504 de-identified outpatient clinical notes, representing 75 patients with at least one primary care visit at KUMC in 2019. Demographic information was included for all notes; however, medication data were not appended as in the MIMIC-III corpus.

#### Data annotation

We adapted medical diagnostic guidelines [20] to establish formal criteria for determining whether there was sufficient information in the clinical note to conclude that patients were probably suffering from insomnia. We considered three categories of information in our criteria: *direct symptoms, indirect symptoms*, and *medications prescribed*. The *direct symptoms* category includes explicit reports of sleep problems, such as persistent concerns about sleep, difficulty initiating sleep at night, or interruptions during sleep. The *indirect symptoms* category encompasses symptoms resulting from disturbances in sleep continuity, such as fatigue or malaise, or daytime sleepiness. The *medications prescribed* category comprises the medications prescribed to treat insomnia. These are classified into primary insomnia medications (exclusively indicated for insomnia, e.g., Estazolam, Flurazepam, Suvorexant, Zolpidem), and secondary insomnia medications (primarily prescribed for other conditions but also beneficial for treating insomnia, e.g., Acamprosate, Diphenhydramine, Gabapentin, Lorazepam). We detail our criterion in the Annotation Guidelines (see the Supplementary Material).

We manually annotated our collection of 185 clinical notes from the MIMIC corpus (see Fig. 1), and assigned one of two labels: “Insomnia” and “Not Insomnia”. Our gold standard dataset includes 101 notes (54.6%) labeled as “Insomnia” and 84 notes (45.4%) as “Not Insomnia”. We divided our gold standard dataset into training (90 notes), validation (24 notes) and test (71 notes) sets, maintaining the same proportion of “Insomnia” and “Not Insomnia” labels across all subsets. To prevent data contamination between the three sets, we ensured that all notes from a single patient were exclusively contained within one set, thereby avoiding the presence of notes from the same patient in multiple sets. Inter-annotator agreement (IAA), measured using F1 score, was 94.6 on the MIMIC corpus. We manually and independently annotated all 504 notes from KUMC using a similar procedure. Among these notes, 53 notes (10.5%) were labeled as “Insomnia” and 451 notes (89.5%) as “Not Insomnia”. The IAA for the KUMC corpus was 88.9 F1 score.

### 2.2 Automatic detection of patients with insomnia

We framed the task of detecting insomnia as a binary classification problem. Our goal was to automatically assign the label “Insomnia” to clinical notes documenting insomnia phenotypes and the label “Not Insomnia” to all other notes, using notes enriched with the patient’s demographic and medication information. We propose a generative approach to perform this task, leveraging the capabilities of state-of-the-art Large Language Models (LLMs). These models introduce an intuitive interface that allows users to describe tasks in natural language and receive responses in kind. Beyond predicting the most likely labels, the models provide explanations for their decisions, adhering to annotation guidelines much like a human annotator would.

To evaluate the effectiveness of our generative approach, we compared its performance to that of a conventional supervised classification method, with BERT-based systems serving as the baseline models. Although LLMs achieve state-of-the-art performance across various NLP tasks, due to their task-specific training, fine-tuned BERT models continue to be strong contenders for information extraction tasks. We describe both approaches below.

### 2.3 BERT-based classification

BERT (Bidirectional Encoder Representations from Transformers) [5] is based on the Transformer architecture. Introduced by Vaswani et al. in 2017 [23], this architecture represents a major innovation in the field of NLP. Unlike previous sequence processing systems that relied heavily on recurrent neural networks (RNNs) [21], the transformer architecture replaces recurrent processes with self-attention mechanisms. While RNNs can also utilize self-attention in combination with recurrence, the transformer architecture uniquely discards recurrent processing altogether in favor of fully leveraging self-attention across the entire sequence. This architecture allows each position in the sequence to attend directly to every other position, thereby capturing complex interdependencies without regard to their distance in the input sequence [23]. This mechanism not only improves the model’s ability to learn contextual relationships between words in text but also significantly reduces training and inference times [16] by enabling greater computational parallelization. The transformer architecture quickly became the foundation for many modern NLP systems, due to its effectiveness and efficiency in handling various tasks such as translation, summarization, and text generation [14].

Among the models based on the transformer architecture, BERT excels in information extraction tasks [5]. Relying on the encoder component of the transformer architecture, BERT utilizes a stack of multi-headed self-attention layers to handle both left and right context of an input sequence simultaneously. This bidirectional nature allows for a comprehensive understanding of the context across the entire sequence, which is critical for tasks requiring nuanced interpretation of text, such as named entity recognition and text classification [14]. Another factor contributing to the success of the BERT model is its compatibility with transfer learning approaches [25]. Transfer learning (TL) is a machine learning method where a model developed for one task is reused as the starting point for a model on a second task. A critical component of TL is pretraining, which involves training a model on a vast amount of data before it is transferred to another task. In the context of NLP, the pretraining process consists of training the model on large text datasets in an unsupervised manner, which equips the model with a rich, generalized understanding of language. Specifically, BERT’s pretraining includes masked language modeling and next sentence prediction tasks [5], which help the model develop a nuanced understanding of language context and relationships.

Originally pretrained on general domain corpora like Books Corpus and Wikipedia [27], BERT can be further pretrained in an unsupervised manner on domain-specific corpora to adapt the model to a particular domain. This approach is particularly effective in the biomedical and clinical domains, where language and terminology significantly diverge from the general text on which BERT was originally trained [15].

However, a fundamental limitation of the original BERT architecture is its maximum input length of 512 tokens. This constraint is particularly problematic in the medical domain, where clinical notes often exceed this limit. In our MIMIC and KUMC corpora, clinical notes contained up to 2,729 and 3,614 words, respectively, well beyond BERT’s capacity. Truncating these documents could lead to loss of critical information, compromising model performance.

To address this issue, we adopted transformer-based variants of BERT that support long input sequences: Longformer [2] and BigBird [26]. These models extend the capabilities of BERT by incorporating efficient attention mechanisms that scale to longer inputs, making them better suited for our task. Specifically, we experimented with four different models:

- **Longformer**: This model extends BERT by incorporating a combination of local windowed attention and task-specific global attention mechanisms [2]. This design allows Longformer to process sequences up to 4,096 tokens efficiently, making it suitable for tasks involving long documents.
- **Clinical-Longformer**: Building upon Longformer, Clinical-Longformer is further pre-trained on 2 million clinical notes extracted from the MIMIC-III database [15].
- **BigBird**: BigBird modifies the attention mechanism of the original BERT by introducing a sparse attention pattern that combines global, random, and local attention [26]. This approach allows the model to handle sequences up to 4,096 tokens with improved efficiency.
- **Clinical-BigBird**: Similar to Clinical-Longformer, Clinical-BigBird is an adaptation of BigBird that has been further pre-trained on 2 million clinical notes extracted from the MIMIC-III database [15].

An additional aspect of TL that greatly benefits BERT is supervised finetuning [25]. Once pretrained, BERT-based models can be further fine-tuned to tackle specific downstream tasks. This process involves training the models on a smaller, task-specific dataset while retaining the general knowledge acquired during the pretraining phase [5]. In this work, we fine-tuned all BERT-based systems to tackle the identification of patients likely to suffer from insomnia, by training the models on a binary text classification task.

### 2.4 Large language models

In the last five years, the field of NLP has been revolutionized by a new family of generative models. These generative models, which include prominent examples such as GPT (Generative Pre-trained Transformer) [4] –the model powering the well-known chatbot ChatGPT– have demonstrated unprecedented capabilities in writing coherent and contextually relevant text using large language models (LLMs). Relying on the decoder component of the transformer architecture, these models generate text by sequentially predicting the next word in a sentence based on previously generated words [23]. This autoregressive approach has made LLMs highly effective in tasks such as text completion, summarization, translation, and even creative writing [14]. Moreover, the extensive size of these models, which for the largest comprising trillions of trainable parameters, has been a key factor in their success [9]. Billions of parameters in these models enable them to capture a rich, nuanced understanding of language patterns and complexities not feasible with smaller models [12].

To harness the potential of these models, prompt engineering has become a crucial technique [17]. The prompt –a set of carefully crafted instructions in natural language– is designed to specify the desired output from the model. The effectiveness of prompt engineering in LLMs is rooted in their training process. Similar to BERT-based models (see Section 2.3), LLMs also benefit from a TL approach. In the case of LLMs, TL typically consists of two major phases: unsupervised pretraining and instruction fine-tuning [17]. During unsupervised pretraining, the models are trained to predict the next word in a sentence given the words that preceded it, thus exposing them to vast amounts of text data. Following pretraining, LLMs often undergo a phase called instruction fine-tuning, where they are specifically trained to follow instructions contained in prompts [4]. In this way, prompt engineering leverages these trained capabilities to guide the model’s output toward solving specific tasks without the need for extensive fine-tuning.

Another aspect that has further expanded the impact of this technology, particularly in high-stakes domains such as medicine, is the advent of open-source LLMs. When handling clinical data, open-source models offer researchers the flexibility to modify and adapt these models to comply with HIPAA privacy requirements [7]. Unlike closed-source models accessed through proprietary application programming interfaces, open-source models can be operated in secure and isolated environments where clinical data is not shared with any third-party organizations [18]. In this work, we developed an LLM-based approach for detecting patients with insomnia. For this purpose, we utilized the Llama-3 family of open-sourced LLMs [6], one of the largest freely available collections of models that deliver performance comparable to proprietary alternatives. Specifically, we employed two of the largest publicly available models at the time of writing this manuscript: Llama 3.1 70B-Instruct, referred to as Llama 70B, and Llama 3.1 405B-Instruct, referred to as Llama 405B.

### 2.5 Generative AI for insomnia detection

We employed prompt engineering techniques to leverage the predictive capabilities of LLMs. Specifically, we integrated few-shot learning [4] with chain-of-thought (CoT) [24] techniques to develop a system that, given a clinical note, not only predicts whether the patient was likely experiencing insomnia during the encounter but also generates a rationale that aligns coherently with its decision and the note’s content. We restructured the criteria outlined in our annotation guidelines (Section 2.1) into a systematic set of rules to guide the LLMs in their classification process and anchor their explanations. This approach emulates the reasoning process employed by human annotators when evaluating each clinical note. We included the rules within the prompt and required the model to assess whether the patient met the specified conditions, providing justifications for its reasoning using the text from the clinical note. In addition, we instructed the model to provide a binary “yes”/”no” judgment for each component of the rule set, indicating whether the corresponding criteria were satisfied based on the clinical note content. Figure 3 illustrates both the prompt provided to the model and the expected output text. We detail each element of the prompt below.

**Figure 3:**
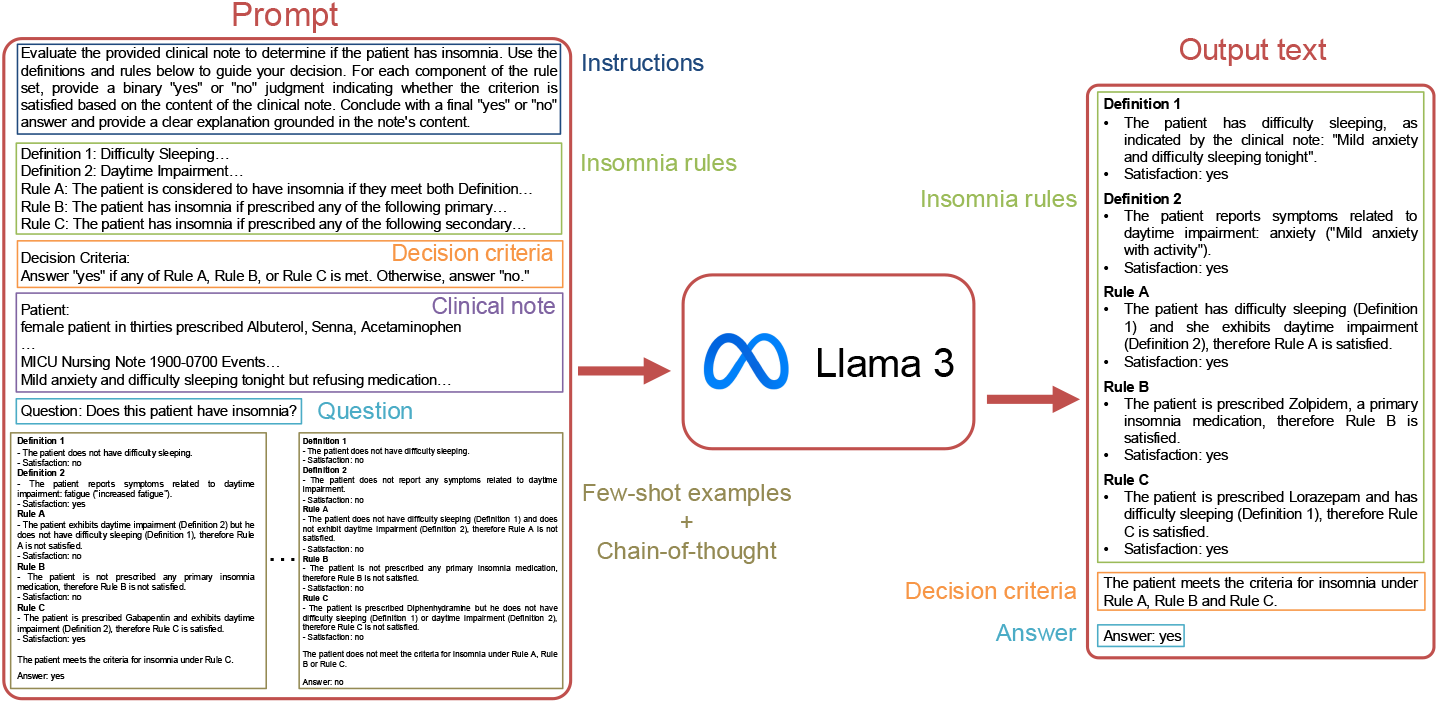
Workflow depicting our LLM-based approach for detecting insomnia.

#### Instructions

A brief set of instructions to resolve the insomnia detection problem. We model the task as a question answering (QA) problem, in which we ask the LLM whether the given patient is likely to have insomnia.

#### Insomnia rules

We reformulated our annotation guidelines into a set of logical rules designed to be more interpretable for the model. These rules comprise two key definitions and three logical implications. As illustrated in Fig. 4, we begin by defining the symptoms associated with difficulty sleeping as *Definition 1* and the symptoms related to daytime impairment as *Definition 2*. Based on these definitions, we outline the following rules:

**Figure 4:**
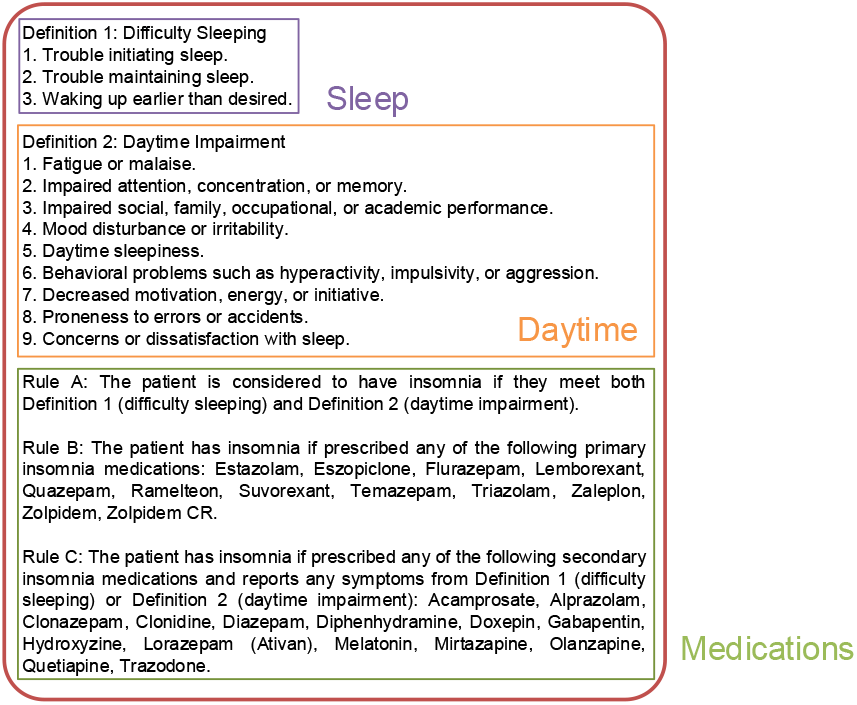
Illustration of the rules used to detect patients likely to suffer from insomnia.

- *Rule A:* A patient is considered likely to be suffering from insomnia if the patient exhibits symptoms from both Definition 1 and Definition 2.
- *Rule B:* A patient is likely to be suffering from insomnia if the patient has been prescribed primary insomnia medications.
- *Rule C:* A patient is likely to be suffering from insomnia if the patient has been prescribed secondary insomnia medications *and* exhibit symptoms from either Definition 1 or Definition 2.

These rules provide a structured framework to guide the model’s classification process.

#### Decision criteria

We inform the model that a patient is considered to suffer from insomnia if they satisfy any of the aforementioned rules.

#### Clinical note

We provide the model with the clinical note, supplemented by the patient’s demographic and medication information.

#### Question

We explicitly ask the model whether the patient suffers from insomnia.

#### Few-shot examples

Few-shot learning is a machine learning framework where a model is designed to learn and make accurate predictions from few training examples [17]. In the context of LLMs, few-shot learning allows these models to leverage their extensive pretraining to tackle new, specialized tasks with just a handful of examples [4]. For our study, we selected eight clinical notes from the training set of the MIMIC corpus as examples—four annotated as “Insomnia” and four as “Not Insomnia” (the eight examples are included in the Supplementary Material). We consistently used these eight examples when applying the models to the MIMIC corpus, and incorporated them and their labels into all prompts. We found that using fewer than eight examples led to decreased performance, while increasing the number of examples beyond eight did not yield any noticeable improvement in the model’s performance on the validation set. When evaluating the models on the KUMC corpus, we adapted this strategy by using a balanced set of eight examples that included four examples from the KUMC corpus and four examples from the original eight selected from the MIMIC training set. As is commonly done, we enhanced the few-shot learning framework with a Chain of Thought (CoT) [24] approach. We demonstrated the logical sequences we followed when solving the eight few-shot examples by incorporating our step-by-step reasoning processes for each example into the prompt. Our observations showed that prompts incorporating few-shot examples, enriched with CoT reasoning, not only improved the transparency of the model’s decisions but also made its outputs more interpretable [19]. In our case, for the four patients likely suffering from insomnia, we extracted the relevant span(s) of text from the clinical notes to show which rule was satisfied and explicitly mentioned the rule in the prompt along with the expected label. For the four patients unlikely to suffer from insomnia, we explained why none of the rules were satisfied before stating the expected label.

### 2.6 Binary Classification Evaluation

We evaluated the binary text classification performance of the competing models using Precision, Recall and F1-score. We call notes labeled “Insomnia” correctly classified by a model true positives (TP) and incorrectly predicted as “Not Insomnia”, false negatives (FN); notes labeled “Not Insomnia” correctly classified true Negative and incorrectly predicted as “Insomnia”, false positives (FP). *Precision* (P) measures the accuracy of positive predictions 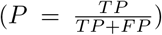; *Recall* (R) measures the ability of the model to identify all positive instances 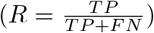; and the *F1-score* (F1) is the harmonic mean of precision and recall, providing a summary of the overall performance of the model 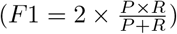.

#### 2.6.1 Explanation Quantitative Evaluation

In addition to evaluate the binary text classification performance of the models, we also assessed the intermediate reasoning steps followed by the generative models when applying the insomnia rules described in Section 2.5. For each clinical note, in addition to assigning a final “Insomnia” or “Not Insomnia” label, our clinical experts also annotated whether the note satisfied each of the five elements of the insomnia rules: Definition 1, Definition 2, Rule A, Rule B, and Rule C. These rule-level annotations were compared against the corresponding rule-level predictions generated by the LLMs during their reasoning process. We treated each rule component as an independent binary classification task, and calculated the P, R and F1 scores to evaluate the performance of the models on each element.

## 3 Results

### 3.1 Binary classification on MIMIC corpus

Table 1 shows the performance of both generative and text classification approaches on our test set from the MIMIC corpus. The highest classification performance is achieved by the Llama 70B model, with a score of 93.0 for precision, recall, and F1. The second-best performance is obtained by the Llama 405B model, with a precision of 95.0, a recall of 88.4, and an F1 score of 91.6. The two generative models outperform the baseline BERT-based models for this binary classification task. In terms of domain-specific BERT-based models, both Clinical-Longformer and Clinical-BigBird outperformed their general-domain counterparts. Clinical-Longformer, with an F1 score of 75.9, surpassed the Longformer model, which had an F1 score of 71.8. Clinical-BigBird, with an F1 score of 81.0, also outperformed its general-domain version, BigBird, which had an F1 score of 80.0.

**Table 1:** Classification results on the MIMIC and KUMC corpora for various BERT-based systems and LLMs. For each metric, the best result is bolded and the second-best is underlined.

| Approach | Model | MIMIC corpus |  |  |  |  |  | KUMC corpus |  |  |
| --- | --- | --- | --- | --- | --- | --- | --- | --- | --- | --- |
|  |  | Test |  |  | Training |  |  |  |  |  |
|  |  | P | R | F1 | P | R | F1 | P | R | F1 |
| Text classification | Longformer | 80.0 | 65.1 | 71.8 | - | - | - | - | - | - |
|  | Clinical-Longformer | 83.3 | 69.8 | 75.9 | - | - | - | - | - | - |
|  | BigBird | 81.0 | 79.1 | 80.0 | - | - | - | - | - | - |
|  | Clinical-BigBird | 82.9 | 79.1 | 81.0 | - | - | - | - | - | - |
| Generative AI | Llama 70B | <u>93.0</u> | **93.0** | **93.0** | <u>95.2</u> | **95.2** | **95.2** | <u>81.8</u> | **86.5** | <u>84.1</u> |
|  | Llama 405B | **95.0** | <u>88.4</u> | <u>91.6</u> | **97.4** | <u>90.5</u> | <u>93.8</u> | **91.3** | 80.8 | **85.7** |

To provide a more comprehensive assessment of the generative approach, we also evaluated it on the training set from our MIMIC corpus. Unlike the BERT-based systems, which were fine-tuned on the training set, our generative approach did not require fine-tuning and used only eight notes from the training set for few-shot learning. Table 1 details the performance of the generative approach on the training set. From the 90 notes in the training subset, we removed the eight notes used as few-shot examples in the prompts (see Section 2.5), using a total of 82 notes for model evaluation. Llama 70B slightly outperformed Llama 405B, with the 70B model achieving a value of 95.2 for precision, recall, and F1 metrics, while the 405B model obtained 97.4, 90.5, and 93.8 for precision, recall, and F1 score, respectively.

### 3.2 Binary classification on KUMC corpus

To assess the generalizability of our generative approach beyond inpatient hospital settings, we evaluated both Llama 70B and Llama 405B on the KUMC corpus. Unlike the MIMIC corpus, which contains ICU discharge summaries and nursing notes, the KUMC dataset comprises outpatient clinical notes from primary care encounters. We excluded the four outpatient notes used as fewshot examples in the prompts (see Section 2.5) from the 504 notes in the KUMC corpus, resulting in 500 notes used for model evaluation. As shown in Table 1, both models maintained strong performance despite the domain shift. Llama 405B achieved the highest F1 score of 85.7, with a precision of 91.3 and recall of 80.8. Llama 70B also performed competitively, achieving an F1 score of 84.1, a precision of 81.8, and a recall of 86.5.

### 3.3 Explanation Quality

We evaluated the rule-level predictions generated by the LLMs during their step-by-step reasoning process described in Section 2.5. Table 2 reports the F1 scores of both Llama 70B and Llama 405B across the five rule components for the MIMIC (training and test sets) and KUMC corpora (see Table S1 in Supplementary Material for an extended evaluation including precision and recall metrics).

**Table 2:** F1 scores for each component of the insomnia rules (Definition 1, Definition 2, Rule A, Rule B, and Rule C) as predicted by the Llama 70B and Llama 405B models. Results are shown for the MIMIC corpus (training and test sets) and the KUMC corpus.

| Corpus |  | Model | Definition 1 | Definition 2 | Rule A | Rule B | Rule C |
| --- | --- | --- | --- | --- | --- | --- | --- |
| MIMIC | Test | Llama 70B | **92.5** | **65.6** | **64.4** | **91.9** | **87.7** |
|  |  | Llama 405B | 90.4 | 64.4 | 58.8 | 88.9 | 80.0 |
|  | Train | Llama 70B | **93.5** | **79.4** | **74.6** | **100.0** | **90.4** |
|  |  | Llama 405B | 90.4 | 72.7 | 67.9 | 97.0 | 88.6 |
| KUMC |  | Llama 70B | 76.9 | 75.5 | 70.8 | 94.5 | **63.4** |
|  |  | Llama 405B | **84.4** | **81.2** | **80.9** | **98.1** | 55.3 |

On the MIMIC test set, Llama 70B outperformed Llama 405B across all rule components. Both models achieved their highest F1 scores on Definition 1, with Llama 70B reaching 92.5 and Llama 405B scoring 90.4. On the MIMIC training set, Llama 70B consistently outperformed Llama 405B on all rule components. Rule B was the most accurately predicted element, where Llama 70B achieved perfect classification (F1 = 100) and Llama 405B reached an F1 score of 97.0.

On the KUMC corpus, Llama 405B achieved higher F1 scores than Llama 70B for all components except Rule C, where Llama 70B outperformed Llama 405B (F1 = 63.4 vs. 55.3). As in the MIMIC training corpus, Rule B yielded the highest F1 scores, with Llama 405B achieving 98.1 and Llama 70B obtaining 94.5.

In contrast, Rule A was generally the most challenging component for both models across datasets. This can be attributed to the models’ difficulty in detecting symptoms associated with Definition 2 (daytime impairment), which consistently yielded lower performance than Definition 1 (difficulty sleeping) across all datasets. Similarly, Rule C posed greater difficulty in the KUMC corpus, where both models exhibited their lowest rule-level performance.

### 3.4 Error Analysis

To better understand the limitations of the generative models, we conducted an error analysis based on the predictions made by the best-performing model for each corpus: Llama 70B for MIMIC and Llama 405B for KUMC. We focused our analysis on the most challenging rule components in each dataset, as identified by the rule-level evaluation: Definition 2 and Rule C in MIMIC, and Definitions 1 and 2 and Rule C in KUMC^1^. Table 3 summarizes representative false positive (FP) and false negative (FN) cases for these components.

**Table 3:**
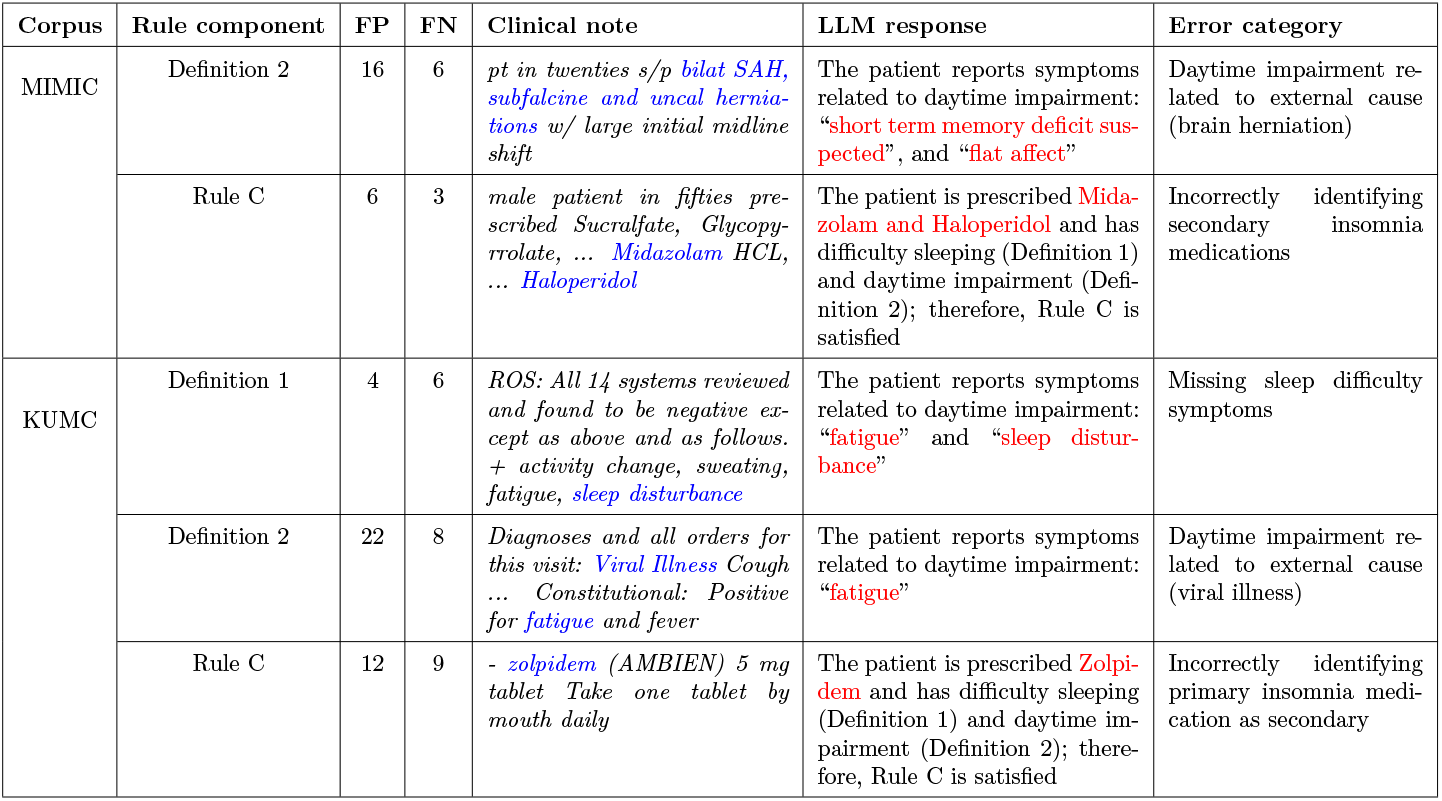
Representative errors made by the best-performing generative model for each corpus (Llama 70B for MIMIC, Llama 405B for KUMC) across the most challenging rule components. FP = false positives, FN = false negatives.

A recurring pattern across both datasets was the difficulty in accurately identifying symptoms related to daytime impairment (Definition 2). This component yielded one of the lowest F1 scores in the rule-level evaluation, and our qualitative analysis confirmed that the LLMs frequently misinterpreted general symptoms such as “anxiety”, “fatigue”, or “memory issues” without adequately considering the broader clinical context. For instance, in the MIMIC corpus, a patient recovering from a traumatic brain injury was incorrectly flagged as experiencing daytime impairment due to cognitive symptoms more likely attributable to the underlying neurological condition. Similarly, in the KUMC corpus, fatigue was often misattributed to insomnia, despite being plausibly linked to other conditions such as viral illness.

Errors in Rule C were also common, particularly when the models misclassified medications as being for insomnia. In the MIMIC corpus, the models incorrectly labeled medications such as Haloperidol and Midazolam—used primarily for agitation or sedation in ICU settings—as secondary insomnia treatments. This issue was even more pronounced in the KUMC corpus, where misclassification of medications accounted for the majority of errors in Rule C, the most challenging component in that dataset. For example, the model erroneously treated medications such as Tramadol, Nortriptyline, and Modafinil as secondary insomnia treatments. Some false positives in the KUMC corpus also resulted from Zolpidem (a primary insomnia medication) being incorrectly treated as a secondary medication, thus incorrectly satisfying Rule C in addition to Rule B.

In the KUMC corpus, the models also made errors on Definition 1 by failing to recognize implicit mentions of sleep difficulty. For example, notes containing vague references to “sleep disturbance” were correctly interpreted as indicators of daytime impairment (Definition 2), but not simultaneously as evidence of difficulty initiating or maintaining sleep (Definition 1). As a result, the models missed several Definition 1 assignments despite the term’s potential applicability to both definitions.

## 4 Discussion

In this study, we present a novel, fully automated system for identifying patients who are likely to be experiencing insomnia by leveraging advanced generative AI models applied to their electronic health records. Our approach demonstrates superior performance over traditional text classification methods, achieving high predictive accuracy on both inpatient and outpatient clinical corpora.

On the MIMIC corpus, both Llama 70B and Llama 405B achieved F1 scores above 90, outperforming the best BERT-based model, Clinical-BigBird, which had an F1 score of 81.0. Other transformer-based baselines performed worse, with general-domain models such as Longformer and BigBird having F1 scores of 71.8 and 80.0, respectively. Our results suggest that while domain adaptation improves the effectiveness of text classifiers for predicting the insomnia status of patients, generative LLMs offer a substantial performance advantage even without the need for task-specific fine-tuning. This underscores the transformative potential of LLMs for robust, scalable phenotyping of complex clinical conditions like insomnia directly from unstructured EHR narratives.

To evaluate the generalizability of our generative approach, we further assessed the performance of the models on the KUMC corpus, which is markedly different from MIMIC in terms of both clinical setting and documentation style. The outpatient notes at KUMC are summary progress notes, generally covering a broad range of systems in a somewhat structured manner, but vary considerably in format and content depending on the provider and clinical team. A key distinction between the two corpora lies in the presence of structured medication data. In the MIMIC corpus, we systematically appended a comprehensive list of medications prescribed during the hospital stay to each clinical note (see Fig. 2), ensuring that the generative models had direct access to this information in a standardized format. In contrast, the KUMC corpus did not include structured medication fields; any mention of medications appeared within the unstructured free text and was inconsistently documented. Consequently, models evaluated on the KUMC data were required to rely more heavily on their natural language understanding capabilities to identify medications and context related to insomnia, without the advantage of explicit structured cues. This scenario more accurately reflects real-world outpatient practice, where medication information is often embedded informally in provider notes and structured fields may be incomplete or absent.

Despite this added complexity, both generative models maintained strong performance on the KUMC corpus. Llama 405B achieved an F1 score of 85.7, while Llama 70B reached 84.1. Notably, the models performance on each corpus approached the inter-annotator agreement (IAA) observed among expert human annotators: 94.6 F1 for MIMIC and 88.9 F1 for KUMC. Since IAA defines an approximate ceiling for achievable model performance on classification tasks like clinical phenotyping, these results suggest that the generative models are nearing human-level accuracy. The slightly lower model performance and IAA on KUMC further emphasize the inherent difficulty of phenotyping from outpatient notes, but also reinforce the effectiveness of our approach in generalizing to real-world clinical settings.

### 4.1 Limitations and future work

While our results demonstrate the effectiveness of generative language models to identify likely insomnia cases using clinical text, several limitations warrant consideration. First, our evaluation was limited to two datasets, that is, MIMIC and KUMC, which differ in setting, level of care, and documentation practices, but may not fully capture the range of documentation practices in other institutions or specialties. Evaluation of additional datasets is needed to confirm generalizability.

Second, error analysis revealed two main challenges: the occasional misclassification of medications as insomnia treatments due to LLM hallucinations [10], and the difficulty distinguishing insomnia symptoms from those attributable to severe underlying disorders, particularly in complex inpatient cases. This was particularly evident in the MIMIC corpus, where patients often presented with critical underlying conditions such as neurological deficits or organ failure. In such cases, their symptoms, such as fatigue or impaired concentrating ability, were mistakenly attributed to insomnia. These findings indicate that current models can sometimes bypass explicit constraints or misinterpret the clinical context.

To mitigate these limitations, future work should explore multi-agent LLM frameworks [8], which is a natural extension of our present approach.

## 5 Conclusion

We introduced a generative AI-based system for identifying patients likely to suffer from insomnia using unstructured clinical notes. Our approach outperformed traditional classification models and achieved performance levels approaching human agreement, demonstrating both accuracy and interpretability. The system generalized well across inpatient and outpatient settings, despite notable differences in the documentation style and structure. Ultimately, our framework offers a scalable and interpretable solution for clinical phenotyping of insomnia and can serve as a blueprint for similar efforts targeting other underdiagnosed or underdocumented conditions in the EHR.

## Acknowledgments

The study was partially funded by the University of Kansas Medical Center Clinical and Translational Science Award grant UL1TR002366 from the National Center for Advancing Translational Sciences.

## Data Availability

We make our generative approach for insomnia phenotyping fully open-source to promote transparency, reproducibility, and further innovation in clinical NLP. All code, prompts, and the manually annotated version of the MIMIC corpus are publicly available at https://github.com/guilopgar/Insomnia-Phenotype-LLM. Access to the original MIMIC-III dataset requires credentialed approval through PhysioNet. De-identified notes from the KUMC corpus will be made available through a controlled access process due to the sensitive nature of outpatient clinical text. Researchers interested in obtaining access must be trained in and compliant with HIPAA regulations. To initiate a data access request, please contact the corresponding author. All requests will be reviewed in accordance with institutional policies, and access will be granted under data use agreements defined by the University of Kansas Medical Center based on its official guidelines.

## Footnotes

1 Although Rule A was the most challenging component on the MIMIC corpus and the second most challenging on the KUMC corpus, we did not include it in this analysis because its errors were fully explained by misclassifications in Definitions 1 and 2, which Rule A depends on.

