## Supplementary Material for "Automated Insomnia Phenotyping from Electronic Health Records: Leveraging Large Language Models to Decode Clinical Narratives"

**APPENDIX 1. List of sleep disturbance terms**

Difficulty in sleep maintenance
Insomnia unspecified
adjustment insomnia
awakening early
awakening early morning
bad dream
bad dreams
bizarre dreams
bothered by difficulty sleeping
broken sleep
cannot get off to sleep
cannot sleep at all
chronic insomnia
complaining of insomnia
complaining of nightmares
complaining of vivid dreams
delayed onset of sleep
difficult sleeping
difficulties sleep
difficulties sleeping
difficulty falling asleep
difficulty getting to sleep
difficulty in sleep initiation
difficulty in sleep maintenance
difficulty sleep
difficulty sleeping
difficulty sleeping through the night
difficulty staying asleep
difficulty staying or falling asleep
disturbance in sleep
disturbance in sleep behaviour
disturbance of sleep behavior
disturbances of sleep
disturbances sleep
disturbed sleep
dysnystaxis
early awakening
early morning awakening
early morning waking
early waking
fitful sleep
frequent night waking
frightening dreams

hard to sleep through night
hard to sleep through the night
have nightmares
hyposomnia
impaired sleep
inability to sleep
initial insomnia
insomnia
insomnia chronic
insomnia late
insomnia matutinal
insomnia vesperal
interrupted sleep
interrupting sleep
keeps waking up
late insomnia
light sleep
light sleeping
lights sleeping
matutinal insomnia
middle insomnia
my sleep was restless
night wake
night wakes
night waking
nightmare
nightmares
not getting enough sleep
organic insomnia
other insomnia
persistent insomnia
poor quality sleep
poor sleep
poor sleep pattern
primary insomnia
problems with sleeping
prolonged sleep
restless sleep
restless sleeping
short of sleep
short sleeping
sleep deficit
sleep deprivation
sleep difficult

sleep difficulties
sleep disorder insomnia
sleep disorder insomnia chronic
sleep disorders
sleep disturbance
sleep disturbances
sleep disturbed
sleep dysfunction
sleep is restless
sleep restless
sleep was restless
sleeping difficulties
sleeping difficulty
sleeping disturbances
sleeplessness
terminal insomnia
terrifying dreams
tosses and turns in sleep
trouble falling asleep
trouble sleeping
unable to sleep
unpleasant dreams
vesperal insomnia
vivid dreams
wakes and cannot sleep again
wakes early
wakes up at night
wakes up during night
waking during night
waking night
waking too early

**APPENDIX 2. List of daytime dysfunction terms**

Aggressive behavior
Altered mood
Anger reaction
Angers
Anxiety
Anxiety NOS
Anxiety disorder
Anxiety state NOS
Anxiety state unspecified
Anxiety state, unspecified
Anxiety states
Anxious behavior
Asthenia
Behavior hyperactive
Behaviour hyperactive
Change in mood
Changeable mood
Complaining of "tired all the time"
Concentration abnormal
Concentration impaired
Concentration impairment
Daytime Sleepiness
Daytime Somnolences
Debility
Decrease in energy
Decrease in motivation
Decreased Mood
Decreased motivation
Depressive disorder NOS
Difficulty controlling anger
Diminished ability to concentrate
Diminished motivation
Disorder of affect
Disturbance of memory
Drowsiness
Dysmnesia
Energy decreased
Enfeebled
Excessive somnolence
Exhaustion, NOS
Fatigue (lassitude)
Fatigue NOS
Fatigued

Feel anxious
Feeling angry
Feeling anxious
Feeling bad
Feeling irritated
Feeling low
Feeling of Nervousness
Feeling of sadness
Feeling sad
Feeling tense
Feeling unhappy
Feeling weak
Feels bad
General inattentiveness
General weakness
Hyperactive behaviour
Impulsive
Increased purposeful goal-directed activity
Increased stress
Instability emotional
Irritability and anger
Irritable
Irritable Moods
LOM - Loss of memory
Labile Moods
Lability emotional
Lack of vitality
Lacking in energy
Lethargy
Loss of motivation
Low mood
Low motivation
Mad
Malaise
Malaise and fatigue
Memory Losses
Memory dysfunction
Memory impaired
Memory impairment
Memory problems
Mental concentration difficult
Mental concentration difficulty
Mental concentration impaired
Mental status, drowsy

Mild anxiety
Mild depression
Minimal depression
Miserable
Mixed anxiety and depressive condition
Mixed anxiety and depressive disease
Mixed anxiety and depressive disorder
Mood changes
Mood disorder, NOS
Mood variable
Mood, Irritable
Moody
More active than typical
Morose mood
Motivational deficit
Motor Hyperactivity
Nervously anxious
Nervousness
Outburst of anger
Outbursts of rage
Physical assaultive behavior
Physical assaultive behaviour
Physically assaultive behavior
Physically assaultive behaviour
Poor concentration
Poor memory
Poor short-term memory
Rapidly tired
Sad
Sadness
Short-term memory poor
Sleepiness
Sleepiness, Daytime
Sometimes tired
Somnolence, Daytime
Somnolent
TATT
TATT - Tired all the time
Tired
Tired all the time
Tired out
Tiredness
Tires quickly
Torpid

Unspecified acute reaction to stress
Unspecified anxiety disorder
Unspecified mood affective disorder
Unstable mood
Variable mood
Washed out
Washed-out
Weakness - general
Weakness generalised
Weakness generalized
absence of motivation
affect unhappy
aggression
aggressive
aggressive behaviors
aggressive behaviour
aggressive behaviours
aggressiveness
altered moods
anger
anger outburst
anger outbursts
angry
angry mood
angst
anxiety
anxiety chronic
anxiety depression mixed
anxiety disorder
anxiety state
anxiety with depression
anxious
anxious mood
anxiousness
asthenic
attitude inattentive
bad memory
behavior aggressive
behavior; overactivity
concentrate inability
concentration difficulties
concentration difficulty
concentration lack
concentration loss

daytime sleepiness
daytime somnolence
debility
decreased energy
decreased interest
demonstrates lethargy
depressed
depressed mood
depression anxiety
depression mild
depression minimal
depression symptom
depression with anxiety
depressions symptoms
depressive disorder
depressive symptoms
difficulty concentration
difficulty managing anger
difficulty remembering
disorder mood
disturbed concentration
disturbed memory
disturbed mental concentration
drowsiness
drowsiness was observed
drowsy
dysmnesia
dysphoria
dysphoric mood
easily tired
easily tires
easily tiring
emotional instability
emotional lability
emotional stress
emotional upset
emotionally labile
energy loss
excessive sleep
excessive sleepiness
excessive sleepiness during day
excessive sleepiness during the day
exhaustion
fatigue

fatigue malaise
fatigues
fatiguing
feel bad
feel tired
feel weak
feeling anxious
feeling down
feeling irritable
feeling nervous
feeling tense
feeling tired
feeling tired all the time
feeling tired or poorly
feeling weak
feeling weakness
feelings of weakness
feels weak
forgetful
forgetfulness
fussiness
general malaise
generalized weakness
having little energy
homicidal tendencies
hyperactive
hyperactive behavior
hyperactivity
impair memory
impaired concentration
impaired memory
impairment memory
impulsiveness
inability to concentrate
inability to maintain attention
inattention
inattentive
irritability
irritable
irritable mood
labile mood
lack concentration
lack of concentration
lack of energy

lack of interest
lack of motivation
lacking energy
lacking motivation
lassitude
lethargic
loss memory short term
loss memory short-term
loss motivation
loss of interest
loss of memory
malaise and fatigue
memory deficit
memory deficits
memory disturbance
memory impairment
memory loss
memory loss or impairment
memory poor
memory problem
memory problems
miserable
mixed anxiety depression
mood angry
mood anxious
mood change
mood disorder
mood disorder nos
mood dysphoric
mood irritable
mood labile
mood lability
mood swing
mood swings
moody
motor hyperactivity
muscle weakness
nervous tension
nervousness
outbursts of anger
overactivity
physical aggression
poor concentration
poor memory

short term memory loss
short-term memory loss
sleepiness
sleepiness daytime
sleepy
somnolence
somnolence excessive
somnolent
tatt
tense
tense feeling
tenseness
tire easily
tired
tired all the time
tired feeling
tiredness
tires easily
tiring easily
trouble concentrating
unable to concentrate
unhappy
unmotivated
violent aggressive behavior
violent behavior
weak
weak feeling
weakness
weariness
worn out

**APPENDIX 3. Insomnia Annotation Guidelines**

Insomnia is a sleep disorder where the patient may have difficulty falling asleep, staying asleep and/or non-restorative sleep leading to daytime dysfunction.

#### Insomnia Definition

According to the International Classification of Sleep Disorders-3$^{rd}$ Edition (ICSD-III), insomnia disorder can be classified as:

1. **Chronic Insomnia Disorder**

   Patient experiences sleep disturbances for the last three months affecting the nighttime sleep for at least three times a week and affects the patients' overall functioning.

2. **Short-term Insomnia Disorder**

   Sleep disturbances experienced for less than three months and affects the patients' overall functioning.

3. **Other Insomnia Disorder**

   Sleep disturbances that do not meet the criteria for chronic insomnia or short-term insomnia disorder are classified under this category.

#### Note Level Annotation

There are several different categories of information that the annotator should consider when labeling a note with Value of "Insomnia". Table 1 provides a listing of categories that should be considered during annotation.

##### *Insomnia Label*

Using these categories (Table 1) as a guideline, the annotator should use their best judgment and domain knowledge when assessing whether a note indicates insomnia.

Table 1.: Categories of text that that would assist in diagnosing insomnia.

| Category | Details | Include | Exclude |
|---|---|---|---|
| Direct Symptoms | Patient reported symptoms | • Difficulty falling asleep at night<br>• Waking up during the night<br>• Waking up too early<br>• Not feeling well-rested after a night's sleep | All other symptom mentions |

| | | | |
|---|---|---|---|
| | | • Daytime tiredness or sleepiness<br>• Irritability, depression or anxiety<br>• Difficulty paying attention, focusing on tasks or remembering<br>• Increased errors or accidents<br>• Ongoing worries about sleep | |
| Indirect Symptoms | Symptoms that are a consequence of the sleep continuity disturbance | • Fatigue or malaise.<br>• Attention, concentration, or memory impairment.<br>• Impaired social, family, occupational, or academic performance.<br>• Mood disturbance or irritability.<br>• Daytime sleepiness.<br>• Behavioral problems such as hyperactivity, impulsivity, and aggression.<br>• Reduced motivation, decreased energy, or lack of initiative.<br>• Proneness for errors or accidents.<br>• Concern about sleep or dissatisfaction with sleep | Other indirect symptom mentions |
| Diagnosis | insomnia disorder diagnosis documented by provider | See list of ICD 9/10 diagnoses in Appendices 4 and 5 | |
| Comorbidities | Comorbid conditions/disorders that may contribute | • Alcoholism<br>• Depression<br>• Anxiety<br>• Hyperlipidemia | |

| | | |
|---|---|---|
| | to (or be caused by) insomnia disorder | |
| Medications | Mentions of medications that are prescribed to treat insomnia (*primary insomnia medications*, first 12 medications from the list) and medications that are beneficial for treating insomnia (*secondary insomnia medications*, the remaining 15 medications from the list) | • Estazolam<br>• Flurazepam<br>• Quazepam<br>• Temazepam<br>• Triazolam<br>• Zolpidem<br>• Zolpidem CR<br>• Zaleplon<br>• Eszopiclone<br>• Ramelteon<br>• Suvorexant<br>• Lemborexant<br>• Doxepin<br>• Trazodone<br>• Quetiapine<br>• Diphenhydramine<br>• Olanzapine<br>• Mirtazapine<br>• Melatonin<br>• Acamprosate<br>• Clonidine<br>• Gabapentin<br>• Hydroxyzine<br>• Alprazolam<br>• Clonazepam<br>• Lorazepam<br>• Diazepam |

#### APPENDIX 4. ICD-9 Codes for Insomnia

| | |
|---|---|
| Insomnia, unspecified | 780.52 |
| Transient disorder of initiating or maintaining sleep | 307.41 |
| Persistent disorder of initiating or maintaining sleep | 307.42 |
| Persistent disorder of initiating or maintaining sleep | 307.42 |
| Lack of adequate sleep | V69.4 |
| Insomnia due to medical condition classified elsewhere | 327.01 |
| Transient disorder of initiating or maintaining sleep | 307.41 |
| Persistent disorder of initiating or maintaining sleep | 307.42 |
| Organic insomnia, unspecified | 327.00 |
| Insomnia due to mental disorder | 327.02 |
| Other organic insomnia | 327.09 |
| insomnia with sleep apnea, unspecified | 780.51 |
| Other sleep disturbances | 780.59 |
| Dysfunctions associated with sleep stages or arousal from sleep | 780.56 |
| Sleep disturbance, unspecified | 780.50 |
| Alcohol induced sleep disorders | 291.82 |
| Drug induced sleep disorders | 292.85 |
| Nonorganic sleep disorder, unspecified | 307.40 |

#### APPENDIX 5. ICD-10 Codes for Insomnia

| | |
|---|---|
| Adjustment insomnia | F51.02 |
| Insomnia due to a medical condition | G47.01 |
| Insomnia due to other mental disorder | F51.05 |
| Insomnia unspecified | G47.00 |
| Other insomnia | G47.09 |
| Other insomnia not due to a substance of known physiological condition | F51.09 |
| Other Sleep Disorders not due to A Substance or Known Physiological Condition | F51.8 |
| Sleep Disorder not due to A Substance or Known Physiological Condition, unspecified | F51.9 |
| Paradoxical insomnia | F51.03 |
| Primary insomnia | F51.01 |
| Psychophysiologic insomnia | F51.04 |
| Alcohol use, unspecified with Alcohol-Induced Sleep Disorder | F10.982 |
| Cocaine Abuse with Cocaine-Induced Sleep Disorder | F14.182 |
| Cocaine Dependence with Cocaine-Induced Sleep Disorder | F14.282 |
| Cocaine use, unspecified with Cocaine-Induced Sleep Disorder | F14.982 |
| Opioid abuse with opioid-induced sleep disorder | F11.182 |
| Opioid dependence with opioid induced sleep disorder | F11.282 |

| | |
|---|---|
| Opioid abuse, unspecified with opioid-induced sleep disorder | F11.982 |
| Other Psychoactive Substance Abuse with Psychoactive Substance-Induced Sleep Disorder | F19.182 |
| Other Psychoactive Substance Dependence with Psychoactive Substance-Induced Sleep Disorder | F19.282 |
| Other Psychoactive Substance use, unspecified with Psychoactive Substance-Induced Sleep Disorder | F19.982 |
| Sleep deprivation | Z72.820 |
| Sedative, Hypnotic or Anxiolytic Dependence with Sedative, Hypnotic or Anxiolytic-Induced Sleep Disorder | F13.282 |
| Sedative, Hypnotic or Anxiolytic use, unspecified with Sedative, Hypnotic or Anxiolytic-Induced Sleep Disorder | F13.982 |
| Sedative, Hypnotic or Anxiolytic use, unspecified with Sedative, Hypnotic or Anxiolytic-Induced Sleep Disorder | F13.982 |
| Insufficient Sleep Syndrome | F51.12 |

**APPENDIX 6. Prompts**

We used structured outputs to guide the generative models (Llama 70B and Llama 405B) to produce well-formed, interpretable predictions when classifying clinical notes as indicative of insomnia or not. Both the MIMIC and KUMC prompts followed the same system message and format, differing only in the few-shot examples used to illustrate the task. All prompts utilized few-shot learning combined with chain-of-thought reasoning to enhance model interpretability and performance.

We enforced a **structured output format** by instructing the models to respond in a strict JSON schema. This schema included fields corresponding to each rule component (Definition 1, Definition 2, Rules A, B, and C), as well as the final insomnia status. Each field required both a binary decision (yes or no) and a corresponding natural language explanation. To ensure syntactic validity, we asked the model to escape internal quotes within the explanations and format the entire response as a valid JSON object.

We provided eight few-shot examples to demonstrate both positive and negative cases of insomnia classification, each fully annotated with explanations and formatted JSON responses. These examples were selected from the MIMIC training set and used consistently for all evaluations on the MIMIC corpus.

For the KUMC corpus, we used a mixed few-shot set composed of four examples from MIMIC and four from KUMC, maintaining the same prompt structure and logic.

<u>MIMIC prompt</u>

{'role': 'system'},

{'content: 'You are an expert clinician trained to identify signs of insomnia from clinical notes, following established diagnostic criteria. Your task is to read a clinical note and return a structured JSON object containing your decisions and justifications for each insomnia-related rule.

You must strictly follow the definitions and medication lists below. Do not use any medication that is not explicitly included.

Definitions:

Definition 1: Difficulty Sleeping

The patient is considered to have difficulty sleeping if they report any of the following:

1. Trouble initiating sleep.
2. Trouble maintaining sleep.
3. Waking up earlier than desired.
4. An explicit mention of insomnia.

Definition 2: Daytime Impairment

The patient is considered to have daytime impairment if they report any of the following:
1. Fatigue or malaise.
2. Impaired attention, concentration, or memory.
3. Impaired social, family, occupational, or academic performance.
4. Mood disturbance or irritability.
5. Daytime sleepiness.
6. Behavioral problems such as hyperactivity, impulsivity, or aggression.
7. Decreased motivation, energy, or initiative.
8. Proneness to errors or accidents.
9. Concerns or dissatisfaction with sleep.
10. An explicit mention of insomnia.

Rules:

Rule A: The patient has insomnia if both Definition 1 and Definition 2 are met.

Rule B: The patient has insomnia if prescribed **any of the following primary insomnia medications**:
Estazolam, Eszopiclone, Flurazepam, Lemborexant, Quazepam, Ramelteon, Suvorexant, Temazepam, Triazolam, Zaleplon, Zolpidem, Zolpidem CR.
- **Important**: Only the medications listed above may be used for Rule B. Do not infer or assume based on other medications.

Rule C: The patient has insomnia if:
- They are prescribed **any of the following secondary insomnia medications**, **AND**
- They exhibit any symptom from Definition 1 or Definition 2.
Valid secondary insomnia medications: Acamprosate, Alprazolam, Clonazepam, Clonidine, Diazepam (Valium), Diphenhydramine, Doxepin, Gabapentin, Hydroxyzine, Lorazepam (Ativan), Melatonin, Mirtazapine, Olanzapine, Quetiapine, Trazodone.
- **Important**: **Only** the above medications may be considered for Rule C. You must not use any unlisted drug, even if it is sedative or commonly used for sleep. Do not rely on external knowledge.

Insomnia status:
Answer "yes" if any of Rule A, Rule B, or Rule C is met. Otherwise, answer "no."

Input Format:
"Patient: [clinical note]
Question: Does this patient have insomnia?"

Output Format:

Return your full answer strictly in valid JSON format using the following template. When citing the text from the clinical notes, escape all internal double quotes (`"`) using a backslash (`\"`) to ensure the output is valid JSON.

"Output in JSON format: {
  "definition_1": {
    "explanation": "[Your detailed justification based on the clinical note]",
    "answer": "[yes or no]"
  },
  "definition_2": {
    "explanation": "[Your detailed justification based on the clinical note]",
    "answer": "[yes or no]"
  },
  "rule_a": {
    "explanation": "[Your justification based on the previous answers for Definition 1, and 2]",
    "answer": "[yes or no]"
  },
  "rule_b": {
    "explanation": "[Your detailed justification based on the clinical note]",
    "answer": "[yes or no]"
  },
  "rule_c": {
    "explanation": "[Your detailed justification based on the clinical note]",
    "answer": "[yes or no]"
  },
  "insomnia_status": {
    "explanation": "[Your justification based on the previous answers for Rule A, B, and C]",
    "answer": "[yes or no]"
  }
}"'},

### Example 1 (‘Insomnia’)

{'role': 'user'},
{'content: [MIMIC note id: 1261284]

Question: Does this patient have insomnia?
Output in JSON format: '},

{'role': 'assistant'},
{'content: '{
  "definition_1": {
    "explanation": "The patient has difficulty sleeping, as indicated by the clinical note: \"he had trouble sleeping\" and \"has not slept in over 3 days\".",
    "answer": "yes"

},
"definition_2": {
"explanation": "The patient reports symptoms related to daytime impairment: fatigue (as indicated by the clinical note: \"He is extremely tired\") and concerns or dissatisfaction with sleep (\"just wants to sleep\").",
"answer": "yes"
},
"rule_a": {
"explanation": "The patient has difficulty sleeping (Definition 1) and exhibits daytime impairment (Definition 2); therefore, Rule A is satisfied.",
"answer": "yes"
},
"rule_b": {
"explanation": "The patient is prescribed Zolpidem, a primary insomnia medication; therefore, Rule B is satisfied.",
"answer": "yes"
},
"rule_c": {
"explanation": "The patient is prescribed Gabapentin and has difficulty sleeping (Definition 1) and daytime impairment (Definition 2); therefore, Rule C is satisfied.",
"answer": "yes"
},
"insomnia_status": {
"explanation": "The patient meets the criteria for insomnia under Rule A, Rule B, and Rule C.",
"answer": "yes"
}
}'},

### Example 2 (‘Not Insomnia’)
{'role': 'user'},
{'content: [MIMIC note id: 11834]

Question: Does this patient have insomnia?
Output in JSON format: '},

{'role': 'assistant'},
{'content: '{
"definition_1": {
"explanation": "The patient does not have difficulty sleeping, as indicated by the clinical note: \"sleeping a lot\".",
"answer": "no"
},
"definition_2": {

"explanation": "The patient does not report any symptoms related to daytime impairment that could likely be attributed to insomnia. The clinical note: \"He reports that for the last 2 weeks, he has worsened unsteadiness and has been very tired, sleeping a lot. He thinks this is due to his XRT treatments which finished 6 days prior to admission\" indicates that the patient's fatigue and daytime sleepiness is due to his cancer treatments, and not insomnia.",
"answer": "no"
},
"rule_a": {
"explanation": "The patient does not have difficulty sleeping (Definition 1) and does not exhibit daytime impairment (Definition 2); therefore, Rule A is not satisfied.",
"answer": "no"
},
"rule_b": {
"explanation": "The patient is not prescribed any primary insomnia medication; therefore, Rule B is not satisfied.",
"answer": "no"
},
"rule_c": {
"explanation": "The patient is prescribed Lorazepam, but he does not have difficulty sleeping (Definition 1) or daytime impairment (Definition 2); therefore Rule C is not satisfied.",
"answer": "no"
},
"insomnia_status": {
"explanation": "The patient does not meet the criteria under Rule A, Rule B, or Rule C.",
"answer": "no"
}
}'},

### Example 3 ('Insomnia')
{'role': 'user'},
{'content: [MIMIC note id: 56727]

Question: Does this patient have insomnia?
Output in JSON format: '},

{'role': 'assistant'},
{'content: '{
"definition_1": {
"explanation": "The patient has difficulty sleeping, as indicated by the clinical note: \"insomnia\".",
"answer": "yes"
},

"definition_2": {
"explanation": "The patient reports symptoms related to daytime impairment, as indicated by the clinical note: \"insomnia\".",
"answer": "yes"
},
"rule_a": {
"explanation": "The patient has difficulty sleeping (Definition 1) and exhibits daytime impairment (Definition 2); therefore, Rule A is satisfied.",
"answer": "yes"
},
"rule_b": {
"explanation": "The patient is prescribed Zolpidem, a primary insomnia medication; therefore, Rule B is satisfied.",
"answer": "yes"
},
"rule_c": {
"explanation": "The patient is prescribed Lorazepam and Clonazepam and has difficulty sleeping (Definition 1) and daytime impairment (Definition 2); therefore, Rule C is satisfied.",
"answer": "yes"
},
"insomnia_status": {
"explanation": "The patient meets the criteria for insomnia under Rule A, Rule B, and Rule C.",
"answer": "yes"
}
}'},

### Example 4 (‘Not Insomnia’)
{'role': 'user'},
{'content: [MIMIC note id: 1267520]

Question: Does this patient have insomnia?
Output in JSON format: '},

{'role': 'assistant'},
{'content: '{
"definition_1": {
"explanation": "The patient does not have difficulty sleeping that could likely be attributed to insomnia. The clinical note: \"he was unable to sleep d/t persistent coughing and bronchospasm\" indicates that the patient’s sleep difficulty is due to his persistent coughing and bronchospasm, and not insomnia.",
"answer": "no"
},

"definition_2": {
"explanation": "The patient reports symptoms related to daytime impairment: fatigue (as indicated by the clinical note: \"extremely fatigued this morning\").",
"answer": "yes"
},
"rule_a": {
"explanation": "The patient exhibits daytime impairment (Definition 2) but does not have difficulty sleeping (Definition 1); therefore, Rule A is not satisfied.",
"answer": "no"
},
"rule_b": {
"explanation": "The patient is not prescribed any primary insomnia medication; therefore, Rule B is not satisfied.",
"answer": "no"
},
"rule_c": {
"explanation": "The patient is not prescribed any secondary insomnia medication; therefore, Rule C is not satisfied.",
"answer": "no"
},
"insomnia_status": {
"explanation": "The patient does not meet the criteria for insomnia under Rule A, Rule B, or Rule C.",
"answer": "no"
}
}'},

### Example 5 (‘Insomnia’)
{'role': 'user'},
{'content: [MIMIC note id: 1277843]

Question: Does this patient have insomnia?
Output in JSON format: '},

{'role': 'assistant'},
{'content: '{
"definition_1": {
"explanation": "The patient has difficulty sleeping, as indicated by the clinical note: \"unable to sleep\" and \"req serax for sleep\".",
"answer": "yes"
},
"definition_2": {
"explanation": "The patient reports symptoms related to daytime impairment, as indicated by the clinical note: \"Slt anxious req serax for sleep, reeval for further needs of

serax\". The need to treat this patient in the hospital with medication (serax) for sleep implies that their inability to sleep is serious enough to cause daytime impairment.",
"answer": "yes"
},
"rule_a": {
"explanation": "The patient has difficulty sleeping (Definition 1) and exhibits daytime impairment (Definition 2); therefore, Rule A is satisfied.",
"answer": "yes"
},
"rule_b": {
"explanation": "The patient is not prescribed any primary insomnia medication; therefore, Rule B is not satisfied.",
"answer": "no"
},
"rule_c": {
"explanation": "The patient is not prescribed any secondary insomnia medication; therefore, Rule C is not satisfied.",
"answer": "no"
},
"insomnia_status": {
"explanation": "The patient meets the criteria for insomnia under Rule A.",
"answer": "yes"
}
}'},

### Example 6 ('Not Insomnia')
{'role': 'user'},
{'content: [MIMIC note id: 1270387]

Question: Does this patient have insomnia?
Output in JSON format: '},

{'role': 'assistant'},
{'content: '{
"definition_1": {
"explanation": "The patient does not have difficulty sleeping.",
"answer": "no"
},
"definition_2": {
"explanation": "The patient reports symptoms related to daytime impairment: low energy (as indicated by the clinical note: \"PT LETHARGIC\").",
"answer": "yes"
},
"rule_a": {

"explanation": "The patient exhibits daytime impairment (Definition 2) but does not have difficulty sleeping (Definition 1); therefore, Rule A is not satisfied.",
"answer": "no"
},
"rule_b": {
"explanation": "The patient is not prescribed any primary insomnia medication; therefore, Rule B is not satisfied.",
"answer": "no"
},
"rule_c": {
"explanation": "The patient is not prescribed any secondary insomnia medication; therefore, Rule C is not satisfied.",
"answer": "no"
},
"insomnia_status": {
"explanation": "The patient does not meet the criteria for insomnia under Rule A, Rule B, or Rule C.",
"answer": "no"
}
}'},

### Example 7 ('Insomnia')
{'role': 'user'},
{'content: [MIMIC note id: 1295870]

Question: Does this patient have insomnia?
Output in JSON format: '},

{'role': 'assistant'},
{'content: '{
"definition_1": {
"explanation": "The patient has difficulty sleeping, as indicated by the clinical note: \"Pt reported unable to sleep overnight\".",
"answer": "yes"
},
"definition_2": {
"explanation": "The patient does not report any symptoms related to daytime impairment that could likely be attributed to insomnia. The clinical note: \"after rec'ing total of MSO4 4mg IVP for back pain during RIJ placement, and sleeping soundly during redtag scan, pt is presently oriented to self only and lethargic\" suggests that the patient is lethargic due to receiving Morphine Sulfate during a procedure rather than from insomnia.",
"answer": "no"
},
"rule_a": {

"explanation": "The patient has difficulty sleeping (Definition 1) but does not exhibit daytime impairment (Definition 2); therefore Rule A is not satisfied.",
"answer": "no"
},
"rule_b": {
"explanation": "The patient is not prescribed any primary insomnia medication; therefore, Rule B is not satisfied.",
"answer": "no"
},
"rule_c": {
"explanation": "The patient is prescribed Lorazepam and has difficulty sleeping (Definition 1); therefore, Rule C is satisfied.",
"answer": "yes"
},
"insomnia_status": {
"explanation": "The patient meets the criteria for insomnia under Rule C.",
"answer": "yes"
}
}'},

### Example 8 (‘Not Insomnia’)
{'role': 'user'},
{'content: [MIMIC note id: 15318]

Question: Does this patient have insomnia?
Output in JSON format: '},

{'role': 'assistant'},
{'content: '{
"definition_1": {
"explanation": "The patient does not have difficulty sleeping.",
"answer": "no"
},
"definition_2": {
"explanation": "The patient does not report any symptoms related to daytime impairment that could likely be attributed to insomnia. The clinical note: \"Abnormal T2 and FLAIR signal within both occipital and parietal lobes in a pattern suggestive of posterior reversible encephalopathy syndrome\" suggests that the patient's symptoms such as \"mild attention problems\" are likely explained by this neurological condition rather than insomnia.",
"answer": "no"
},
"rule_a": {

"explanation": "The patient does not have difficulty sleeping (Definition 1) and does not exhibit daytime impairment (Definition 2); therefore, Rule A is not satisfied.",
"answer": "no"
},
"rule_b": {
"explanation": "The patient is not prescribed any primary insomnia medication; therefore, Rule B is not satisfied.",
"answer": "no"
},
"rule_c": {
"explanation": "The patient is prescribed Clonidine and Lorazepam but does not have difficulty sleeping (Definition 1) or daytime impairment (Definition 2), therefore Rule C is not satisfied.",
"answer": "no"
},
"insomnia_status": {
"explanation": "The patient does not meet the criteria for insomnia under Rule A, Rule B, or Rule C.",
"answer": "no"
}
}'}

KUMC prompt
{'role': 'system'},
{'content: 'You are an expert clinician trained to identify signs of insomnia from clinical notes, following established diagnostic criteria. Your task is to read a clinical note and return a structured JSON object containing your decisions and justifications for each insomnia-related rule.

You must strictly follow the definitions and medication lists below. Do not use any medication that is not explicitly included.

Definitions:

Definition 1: Difficulty Sleeping
The patient is considered to have difficulty sleeping if they report any of the following:
1. Trouble initiating sleep.
2. Trouble maintaining sleep.
3. Waking up earlier than desired.
4. An explicit mention of insomnia.

Definition 2: Daytime Impairment
The patient is considered to have daytime impairment if they report any of the following:
1. Fatigue or malaise.

2. Impaired attention, concentration, or memory.
3. Impaired social, family, occupational, or academic performance.
4. Mood disturbance or irritability.
5. Daytime sleepiness.
6. Behavioral problems such as hyperactivity, impulsivity, or aggression.
7. Decreased motivation, energy, or initiative.
8. Proneness to errors or accidents.
9. Concerns or dissatisfaction with sleep.
10. An explicit mention of insomnia.

Rules:

Rule A: The patient has insomnia if both Definition 1 and Definition 2 are met.

Rule B: The patient has insomnia if prescribed **any of the following primary insomnia medications**:
Estazolam, Eszopiclone, Flurazepam, Lemborexant, Quazepam, Ramelteon, Suvorexant, Temazepam, Triazolam, Zaleplon, Zolpidem, Zolpidem CR.
- **Important**: Only the medications listed above may be used for Rule B. Do not infer or assume based on other medications.

Rule C: The patient has insomnia if:
- They are prescribed **any of the following secondary insomnia medications**, **AND**
- They exhibit any symptom from Definition 1 or Definition 2.

Valid secondary insomnia medications: Acamprosate, Alprazolam, Clonazepam, Clonidine, Diazepam (Valium), Diphenhydramine, Doxepin, Gabapentin, Hydroxyzine, Lorazepam (Ativan), Melatonin, Mirtazapine, Olanzapine, Quetiapine, Trazodone.
- **Important**: **Only** the above medications may be considered for Rule C. You must not use any unlisted drug, even if it is sedative or commonly used for sleep. Do not rely on external knowledge.

Insomnia status:
Answer "yes" if any of Rule A, Rule B, or Rule C is met. Otherwise, answer "no."

Input Format:
"Patient: [clinical note]
Question: Does this patient have insomnia?"

Output Format:
Return your full answer strictly in valid JSON format using the following template. When citing the text from the clinical notes, escape all internal double quotes (`"`) using a backslash (`\"`) to ensure the output is valid JSON.
"Output in JSON format: {

"definition_1": {
"explanation": "[Your detailed justification based on the clinical note]",
"answer": "[yes or no]"
},
"definition_2": {
"explanation": "[Your detailed justification based on the clinical note]",
"answer": "[yes or no]"
},
"rule_a": {
"explanation": "[Your justification based on the previous answers for Definition 1, and 2]",
"answer": "[yes or no]"
},
"rule_b": {
"explanation": "[Your detailed justification based on the clinical note]",
"answer": "[yes or no]"
},
"rule_c": {
"explanation": "[Your detailed justification based on the clinical note]",
"answer": "[yes or no]"
},
"insomnia_status": {
"explanation": "[Your justification based on the previous answers for Rule A, B, and C]",
"answer": "[yes or no]"
}
}"'},

### Example 1 (‘Not Insomnia’)
{'role': 'user'},
{'content: [KUMC note 1]

Question: Does this patient have insomnia?
Output in JSON format: '},

{'role': 'assistant'},
{'content: '{
"definition_1": {
"explanation": "The patient does not have difficulty sleeping.",
"answer": "no"
},
"definition_2": {
"explanation": "The patient does not report any symptoms related to daytime impairment.",
"answer": "no"

},
"rule_a": {
"explanation": "The patient does not have difficulty sleeping (Definition 1) and does not exhibit daytime impairment (Definition 2); therefore, Rule A is not satisfied.",
"answer": "no"
},
"rule_b": {
"explanation": "The patient is not prescribed any primary insomnia medication; therefore, Rule B is not satisfied.",
"answer": "no"
},
"rule_c": {
"explanation": "The patient is prescribed gabapentin, but he does not have difficulty sleeping (Definition 1) or daytime impairment (Definition 2); therefore, Rule C is not satisfied.",
"answer": "no"
},
"insomnia_status": {
"explanation": "The patient does not meet the criteria for insomnia under Rule A, Rule B or Rule C.",
"answer": "no"
}
}'},

### Example 2 ('Insomnia')
{'role': 'user'},
{'content: [KUMC note 2]

Question: Does this patient have insomnia?
Output in JSON format: '},

{'role': 'assistant'},
{'content: '{
"definition_1": {
"explanation": "The patient has difficulty sleeping, as indicated by the clinical note: \"She is having a hard time staying asleep at night. She usually fall asleep fairly easily but seems to wake up middle of the night and most nights is hard for her to go back to sleep\", and \"Insomnia, unspecified type\".",
"answer": "yes"
},
"definition_2": {
"explanation": "The patient reports symptoms related to daytime impairment: concerns or dissatisfaction with sleep (as indicated by the clinical note: \"patient is here by herself

```
for follow up of her blood pressure, edema, and problems with sleep\", and \"She is
interested in trying something to help with sleep\"), and \"Insomnia, unspecified type\".",
      "answer": "yes"
   },
   "rule_a": {
      "explanation": "The patient has difficulty sleeping (Definition 1) and exhibits daytime
impairment (Definition 2); therefore, Rule A is satisfied.",
      "answer": "yes"
   },
   "rule_b": {
      "explanation": "The patient is not prescribed any primary insomnia medication;
therefore, Rule B is not satisfied.",
      "answer": "no"
   },
   "rule_c": {
      "explanation": "The patient is prescribed melatonin and has difficulty sleeping
(Definition 1) and daytime impairment (Definition 2); therefore, Rule C is satisfied.",
      "answer": "yes"
   },
   "insomnia_status": {
      "explanation": "The patient meets the criteria for insomnia under Rule A and Rule C.",
      "answer": "yes"
   }
}'},

# Example 3 ('Not Insomnia')
{'role': 'user'},
{'content: [KUMC note 3]

Question: Does this patient have insomnia?
Output in JSON format: '},

{'role': 'assistant'},
{'content: '{
   "definition_1": {
      "explanation": " The patient does not have difficulty sleeping.",
      "answer": "no"
   },
   "definition_2": {
      "explanation": "The patient does not report any symptoms related to daytime
impairment.",
      "answer": "no"
   },
   "rule_a": {
```

"explanation": "The patient does not have difficulty sleeping (Definition 1) and does not exhibit daytime impairment (Definition 2); therefore, Rule A is not satisfied.",
"answer": "no"
},
"rule_b": {
"explanation": "The patient is not prescribed any primary insomnia medication; therefore, Rule B is not satisfied.",
"answer": "no"
},
"rule_c": {
"explanation": "The patient is not prescribed any secondary insomnia medication; therefore, Rule C is not satisfied.",
"answer": "no"
},
"insomnia_status": {
"explanation": "The patient does not meet the criteria for insomnia under Rule A, Rule B or Rule C.",
"answer": "no"
}
}'},

### Example 4 ('Not Insomnia')
{'role': 'user'},
{'content: [KUMC note 4]

Question: Does this patient have insomnia?
Output in JSON format: '},

{'role': 'assistant'},
{'content: '{
"definition_1": {
"explanation": "The patient does not have difficulty sleeping.",
"answer": "no"
},
"definition_2": {
"explanation": "The patient does not report any symptoms related to daytime impairment.",
"answer": "no"
},
"rule_a": {
"explanation": "The patient does not have difficulty sleeping (Definition 1) and does not exhibit daytime impairment (Definition 2); therefore, Rule A is not satisfied.",
"answer": "no"
},

"rule_b": {
"explanation": "The patient is not prescribed any primary insomnia medication; therefore, Rule B is not satisfied.",
"answer": "no"
},
"rule_c": {
"explanation": "The patient is not prescribed any secondary insomnia medication; therefore, Rule C is not satisfied.",
"answer": "no"
},
"insomnia_status": {
"explanation": "The patient does not meet the criteria for insomnia under Rule A, Rule B or Rule C.",
"answer": "no"
}
}'},

### Example 5 (‘Insomnia’)
{'role': 'user'},
{'content: [MIMIC note id: 1261284]

Question: Does this patient have insomnia?
Output in JSON format: '},

{'role': 'assistant'},
{'content: '{
"definition_1": {
"explanation": "The patient has difficulty sleeping, as indicated by the clinical note: \"he had trouble sleeping\" and \"has not slept in over 3 days\".",
"answer": "yes"
},
"definition_2": {
"explanation": "The patient reports symptoms related to daytime impairment: fatigue (as indicated by the clinical note: \"He is extremely tired\") and concerns or dissatisfaction with sleep (\"just wants to sleep\").",
"answer": "yes"
},
"rule_a": {
"explanation": "The patient has difficulty sleeping (Definition 1) and exhibits daytime impairment (Definition 2); therefore, Rule A is satisfied.",
"answer": "yes"
},
"rule_b": {

"explanation": "The patient is prescribed Zolpidem, a primary insomnia medication; therefore, Rule B is satisfied.",
"answer": "yes"
},
"rule_c": {
"explanation": "The patient is prescribed Gabapentin and has difficulty sleeping (Definition 1) and daytime impairment (Definition 2); therefore, Rule C is satisfied.",
"answer": "yes"
},
"insomnia_status": {
"explanation": "The patient meets the criteria for insomnia under Rule A, Rule B, and Rule C.",
"answer": "yes"
}
}'},

### Example 6 ('Not Insomnia')
{'role': 'user'},
{'content: [MIMIC note id: 11834]

Question: Does this patient have insomnia?
Output in JSON format: '},

{'role': 'assistant'},
{'content: '{
"definition_1": {
"explanation": "The patient does not have difficulty sleeping, as indicated by the clinical note: \"sleeping a lot\".",
"answer": "no"
},
"definition_2": {
"explanation": "The patient does not report any symptoms related to daytime impairment that could likely be attributed to insomnia. The clinical note: \"He reports that for the last 2 weeks, he has worsened unsteadiness and has been very tired, sleeping a lot. He thinks this is due to his XRT treatments which finished 6 days prior to admission\" indicates that the patient's fatigue and daytime sleepiness is due to his cancer treatments, and not insomnia.",
"answer": "no"
},
"rule_a": {
"explanation": "The patient does not have difficulty sleeping (Definition 1) and does not exhibit daytime impairment (Definition 2); therefore, Rule A is not satisfied.",
"answer": "no"

},
"rule_b": {
"explanation": "The patient is not prescribed any primary insomnia medication; therefore, Rule B is not satisfied.",
"answer": "no"
},
"rule_c": {
"explanation": "The patient is prescribed Lorazepam, but he does not have difficulty sleeping (Definition 1) or daytime impairment (Definition 2); therefore, Rule C is not satisfied.",
"answer": "no"
},
"insomnia_status": {
"explanation": "The patient does not meet the criteria under Rule A, Rule B, or Rule C.",
"answer": "no"
}
}'},

### Example 7 ('Insomnia')
{'role': 'user'},
{'content: [MIMIC note id: 56727]

Question: Does this patient have insomnia?
Output in JSON format: '},

{'role': 'assistant'},
{'content: '{
"definition_1": {
"explanation": "The patient has difficulty sleeping, as indicated by the clinical note: \"insomnia\".",
"answer": "yes"
},
"definition_2": {
"explanation": "The patient reports symptoms related to daytime impairment, as indicated by the clinical note: \"insomnia\".",
"answer": "yes"
},
"rule_a": {
"explanation": "The patient has difficulty sleeping (Definition 1) and exhibits daytime impairment (Definition 2); therefore, Rule A is satisfied.",
"answer": "yes"
},
"rule_b": {

"explanation": "The patient is prescribed Zolpidem, a primary insomnia medication; therefore, Rule B is satisfied.",
"answer": "yes"
},
"rule_c": {
"explanation": "The patient is prescribed Lorazepam and Clonazepam and has difficulty sleeping (Definition 1) and daytime impairment (Definition 2); therefore, Rule C is satisfied.",
"answer": "yes"
},
"insomnia_status": {
"explanation": "The patient meets the criteria for insomnia under Rule A, Rule B, and Rule C.",
"answer": "yes"
}
}'},

### Example 8 (‘Not Insomnia’)
{'role': 'user'},
{'content: [MIMIC note id: 1267520]

Question: Does this patient have insomnia?
Output in JSON format: '},

{'role': 'assistant'},
{'content: '{
"definition_1": {
"explanation": "The patient does not have difficulty sleeping that could likely be attributed to insomnia. The clinical note: \"he was unable to sleep d/t persistent coughing and bronchospasm\" indicates that the patient’s sleep difficulty is due to his persistent coughing and bronchospasm, and not insomnia.",
"answer": "no"
},
"definition_2": {
"explanation": "The patient reports symptoms related to daytime impairment: fatigue (as indicated by the clinical note: \"extremely fatigued this morning\").",
"answer": "yes"
},
"rule_a": {
"explanation": "The patient exhibits daytime impairment (Definition 2) but does not have difficulty sleeping (Definition 1); therefore, Rule A is not satisfied.",
"answer": "no"
},
"rule_b": {

"explanation": "The patient is not prescribed any primary insomnia medication; therefore, Rule B is not satisfied.",
"answer": "no"
},
"rule_c": {
"explanation": "The patient is not prescribed any secondary insomnia medication; therefore, Rule C is not satisfied.",
"answer": "no"
},
"insomnia_status": {
"explanation": "The patient does not meet the criteria for insomnia under Rule A, Rule B, or Rule C.",
"answer": "no"
}
}'}

**Table S1. Extended Explanation Quality Evaluation.** Precision (P), Recall (R) and F1 scores (F1) for each component of the insomnia rules (Definition 1, Definition 2, Rule A, Rule B, and Rule C) as predicted by the Llama 70B and Llama 405B models. Results are shown for the MIMIC corpus (training and test sets) and the KUMC corpus.

| **Corpus** | | **Model** | **Definition 1** | | | **Definition 2** | | | **Rule A** | | | **Rule B** | | | **Rule C** | | |
|---|---|---|---|---|---|---|---|---|---|---|---|---|---|---|---|---|---|
| | | | **P** | **R** | **F1** | **P** | **R** | **F1** | **P** | **R** | **F1** | **P** | **R** | **F1** | **P** | **R** | **F1** |
| MIMIC | Test | Llama 70B | 86.0 | 100 | 92.5 | 56.8 | 77.8 | 65.6 | 52.8 | 82.6 | 64.4 | 100 | 85.0 | 91.9 | 84.2 | 91.4 | 87.7 |
| | | Llama 405B | 91.7 | 89.2 | 90.4 | 59.4 | 70.4 | 64.4 | 53.6 | 65.2 | 58.8 | 100 | 80.0 | 88.9 | 80.0 | 80.0 | 80.0 |
| | Train | Llama 70B | 90.0 | 97.3 | 93.5 | 73.0 | 87.1 | 79.4 | 64.7 | 88.0 | 74.6 | 100 | 100 | 100 | 89.2 | 91.7 | 90.4 |
| | | Llama 405B | 91.7 | 89.2 | 90.4 | 68.6 | 77.4 | 72.7 | 61.3 | 76.0 | 67.9 | 100 | 94.1 | 97.0 | 91.2 | 86.1 | 88.6 |
| KUMC | | Llama 70B | 66.7 | 90.9 | 76.9 | 73.1 | 78.1 | 75.5 | 57.5 | 92.0 | 70.8 | 92.9 | 96.3 | 94.5 | 68.4 | 59.1 | 63.4 |
| | | Llama 405B | 87.1 | 81.8 | 84.4 | 74.7 | 89.0 | 81.2 | 86.4 | 76.0 | 80.9 | 100 | 96.3 | 98.1 | 52.0 | 59.1 | 55.3 |
